# Methylation scores for smoking, alcohol consumption, and body mass index and risk of seven types of cancer

**DOI:** 10.1101/2021.02.08.21251370

**Authors:** Pierre-Antoine Dugué, Chenglong Yu, Allison M Hodge, Ee Ming Wong, JiHoon E Joo, Chol-Hee Jung, Daniel Schmidt, Enes Makalic, Daniel D Buchanan, Gianluca Severi, Dallas R English, John L Hopper, Roger L Milne, Graham G Giles, Melissa C Southey

**Author notes:** **Correspondence and requests for reprints:** Pierre-Antoine Dugué, Precision Medicine, School of Clinical Sciences at Monash Health, Monash University, 246 Clayton Rd, Clayton, VIC 3168, Australia. **Novelty & Impact Statements:** Methylation marks of exposure to health risk factors may better capture current and past exposures than questionnaires, and reflect different individual responses to exposure. We calculated several methylation scores for smoking, body mass index and alcohol consumption, using summary statistics from published epigenome-wide association studies, and found that these were associated with cancer risk, independently of health-related confounders. Methylation scores for lifestyle may provide some improvements in the prediction of cancer risk.

## Abstract

Methylation marks of exposure to health risk factors may be useful markers of cancer risk as they might better capture current and past exposures than questionnaires, and reflect different individual responses to exposure. We used data from seven case-control studies nested within the Melbourne Collaborative Cohort Study of blood DNA methylation and risk of colorectal, gastric, kidney, lung, prostate and urothelial cancer, and B-cell lymphoma (N cases=3,123). Methylation scores (MS) for smoking, body mass index (BMI), and alcohol consumption were calculated based on published data as weighted averages of methylation values. Rate ratios (RR) and 95% confidence intervals for association with cancer risk were estimated using conditional logistic regression and expressed per standard deviation increase of the MS, with and without adjustment for health-related confounders. The contribution of MS to discriminate cases from controls was evaluated using the area under the curve (AUC). After confounder adjustment, we observed: large associations (RR∼1.5-1.7) with lung cancer risk for smoking MS; moderate associations (RR∼1.2-1.3) with urothelial cancer risk for smoking MS and with mature B-cell neoplasm risk for BMI and alcohol MS; moderate to small associations (RR∼1.1-1.2) for BMI and alcohol MS with several cancer types and cancer overall. Generally small AUC increases were observed after inclusion of several MS in the same model (colorectal, gastric, kidney, urothelial cancers: +3%; lung cancer: +7%; B-cell neoplasms: +8%). Methylation scores for smoking, BMI, and alcohol consumption show independent associations with cancer risk, and may provide some improvements in risk prediction.

## INTRODUCTION

Population-based estimates indicate that smoking, alcohol consumption and obesity combined explain approximately 20% of cancers diagnosed in Australia.^**1**^ These estimates are based on relatively strong assumptions that health risk factors are measured without error in cohort studies, and that population-average estimates reflect individual-level risk. In recent years, a growing body of research has reported associations of genetic and epigenetic measures with these lifestyle factors, raising the potential to increase our understanding of the role they play in the aetiology of cancer at the individual level. Recent research has made successful use of DNA methylation to study, for example, biological ageing ^**2**, **3**^ and its association with cancer risk and mortality.^**4-8**^

Tobacco smoking, adiposity and alcohol consumption are the health risk factors for which most convincing evidence exist regarding their widespread impact on the methylome; studies have identified thousands of CpG sites at which blood DNA methylation is associated with these exposures.^**9-14**^ This raises potential for increasing the precision with which we measure these health risk factors at the individual level, as it is plausible that i) methylation changes are more abundant and marked in individuals who are more sensitive to these exposures, ii) the levels of exposure collected via cohort questionnaires may be prone to substantial measurement error, and iii) harmful levels of exposure accumulate over the life course and cannot be tracked by only one, two, or more time-point assessments. These aspects likely result in underestimates of association of lifestyle factors with cancer risk. The usefulness of integrated genomic measures to enhance epidemiological exposure assessment is increasingly recognised.^**15-18**^ Recent studies have developed and validated epigenetic predictors for several lifestyle factors.^**11**, **19**^

In this study, we aimed to assess the association between several methylation-based scores for smoking, alcohol consumption and body mass index (BMI) and the risk of cancer in adulthood. We used data from seven prospective case-control studies nested in the Melbourne Collaborative Cohort Study (MCCS), including 3,123 incident cases of colorectal, gastric, kidney, lung, prostate and urothelial cancers and B-cell lymphoma.

## MATERIALS AND METHODS

### Study samples and blood collection

The MCCS is a prospective study of 41,513 adult volunteers (24,469 women) aged between 27 and 76 years (99% aged 40-69) when recruited between 1990 and 1994. More details on the cohort are available elsewhere.^**20**, **21**^

We used data from case-control studies of DNA methylation and risk of colorectal, gastric, kidney, lung, mature B-cell lymphoma, prostate and urothelial cancer, nested within the MCCS.^**22-27**^ These cancers account for about 50% of new cancer cases in Victoria in 2018.^**28**^ DNA was extracted from peripheral blood taken at the time of recruitment (1990-1994) except for 151 case-control pairs of the urothelial cancer study, for which blood samples were taken at a follow-up visit in 2003-2007. The DNA source was dried blood spots, peripheral blood mononuclear cells or buffy coats for 70%, 28% and 2% of participants, respectively. More details can be found elsewhere.^**21**, **29**^ Incident cancer cases were identified by linkage with the Victorian Cancer Registry and the Australian Cancer Database (Australian Institute of Health and Welfare), which are considered to be virtually complete. Cases were followed from inclusion until 2012-2013, so the follow-up time was up to 23 years. The median from recruitment to diagnosis was 9.7 years (interquartile range: 5.4 to 13.6); these figures for individual cancer types are shown in **Supplementary Table 1**. For each nested case-control study, controls were individually matched to incident cases (diagnosed after blood sample collection) on age using incidence density sampling (i.e. they had to be free of the cancer of interest up to the age at diagnosis of the corresponding case), sex, country of birth (Australia/New-Zealand, Southern Europe, Northern Europe), blood DNA source (dried blood spots, peripheral blood mononuclear cells or buffy coat) and blood collection period (baseline or wave 2 for the urothelial cancer study). Controls were also matched to cases on year of birth, except for the colorectal cancer study where controls were matched on year of baseline attendance. For the lung cancer study, controls were also matched on smoking status at the time of blood collection.

**Table 1.**
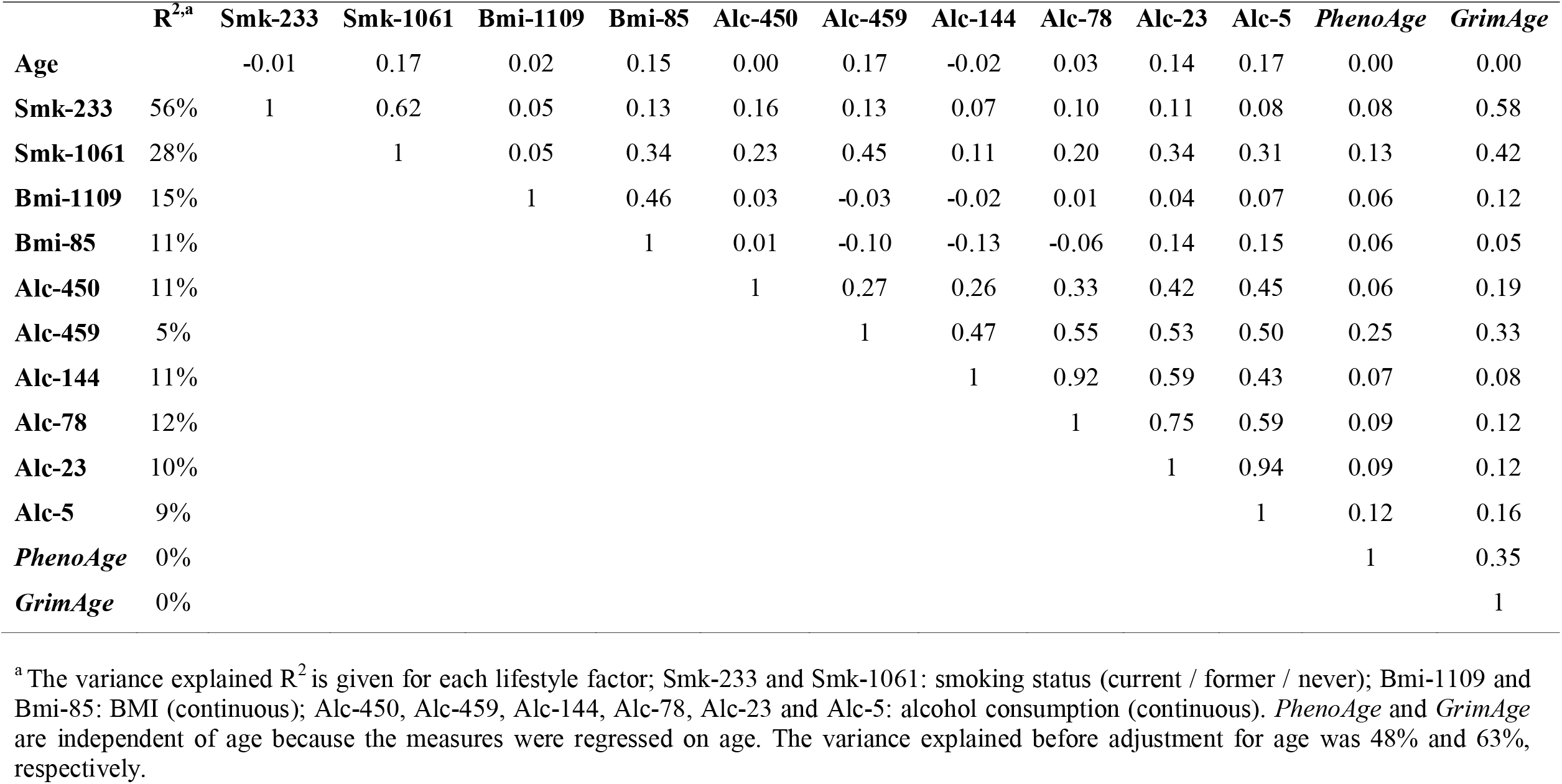
Correlations between methylation scores

|  | **R<sup>2,a</sup>** | **Smk-233** | **Smk-1061** | **Bmi-1109** | **Bmi-85** | **Alc-450** | **Alc-459** | **Alc-144** | **Alc-78** | **Alc-23** | **Alc-5** | **PhenoAge** | **GrimAge** |
| --- | --- | --- | --- | --- | --- | --- | --- | --- | --- | --- | --- | --- | --- |
| **Age** |  | -0.01 | 0.17 | 0.02 | 0.15 | 0.00 | 0.17 | -0.02 | 0.03 | 0.14 | 0.17 | 0.00 | 0.00 |
| **Smk-233** | 56% | 1 | 0.62 | 0.05 | 0.13 | 0.16 | 0.13 | 0.07 | 0.10 | 0.11 | 0.08 | 0.08 | 0.58 |
| **Smk-1061** | 28% |  | 1 | 0.05 | 0.34 | 0.23 | 0.45 | 0.11 | 0.20 | 0.34 | 0.31 | 0.13 | 0.42 |
| **Bmi-1109** | 15% |  |  | 1 | 0.46 | 0.03 | -0.03 | -0.02 | 0.01 | 0.04 | 0.07 | 0.06 | 0.12 |
| **Bmi-85** | 11% |  |  |  | 1 | 0.01 | -0.10 | -0.13 | -0.06 | 0.14 | 0.15 | 0.06 | 0.05 |
| **Alc-450** | 11% |  |  |  |  | 1 | 0.27 | 0.26 | 0.33 | 0.42 | 0.45 | 0.06 | 0.19 |
| **Alc-459** | 5% |  |  |  |  |  | 1 | 0.47 | 0.55 | 0.53 | 0.50 | 0.25 | 0.33 |
| **Alc-144** | 11% |  |  |  |  |  |  | 1 | 0.92 | 0.59 | 0.43 | 0.07 | 0.08 |
| **Alc-78** | 12% |  |  |  |  |  |  |  | 1 | 0.75 | 0.59 | 0.09 | 0.12 |
| **Alc-23** | 10% |  |  |  |  |  |  |  |  | 1 | 0.94 | 0.09 | 0.12 |
| **Alc-5** | 9% |  |  |  |  |  |  |  |  |  | 1 | 0.12 | 0.16 |
| **PhenoAge** | 0% |  |  |  |  |  |  |  |  |  |  | 1 | 0.35 |
| **GrimAge** | 0% |  |  |  |  |  |  |  |  |  |  |  | 1 |

<sup>a</sup>The variance explained $R^2$ is given for each lifestyle factor; Smk-233 and Smk-1061: smoking status (current / former / never); Bmi-1109 and Bmi-85: BMI (continuous); Alc-450, Alc-459, Alc-144, Alc-78, Alc-23 and Alc-5: alcohol consumption (continuous). *PhenoAge* and *GrimAge* are independent of age because the measures were regressed on age. The variance explained before adjustment for age was 48% and 63%, respectively.

### DNA extraction and bisulfite conversion, and DNA methylation data processing

Methods relating to DNA extraction and bisulfite conversion, and DNA methylation data processing have been described previously ^**4**, **29**^ and are detailed in **Supplementary Methods**.

### Methylation scores for smoking, alcohol consumption and BMI

We calculated several methylation scores, all as weighted averages: for smoking we calculated **Smk-233**: score comprising 233 CpGs resulting from the Lasso regression applied to predict smoking pack-years (log-transformed), using as weights the regression coefficients available from the original publication from McCartney et al.^**19**^ and **Smk-1**,**061**: score comprising 1,061 CpGs at which methylation associated with a comprehensive smoking index in the MCCS (P<10^−7^), and also reported to be associated with smoking (P<10^−7^) in previous epigenome-wide association studies (EWAS), using as weights the individual association coefficients from the MCCS EWAS.^**12**^ Similarly, for BMI and alcohol consumption, we calculated, **Bmi-1109** and **Alc-450**, respectively, based on the predictors developed by McCartney et al.; and **Bmi-85** and **Alc-459** based on associations reported with P<10^−7^ in the MCCS and external data.^**13**, **14**^ For alcohol consumption, we also calculated four scores using the weights available from the meta-analysis of Liu et al.^**11**^ These scores included 5, 23, 78 and 144 CpGs (A**lc-5, Alc-23**, A**lc-78** and **Alc-144**) and were reported to explain from 5 to 14% of variance in alcohol consumption in testing datasets. The scores calculated from the studies by McCartney et al. and Liu et al. were obtained via *Lasso* and *Elastic net* regression, respectively, hence they aimed to predict these lifestyle factors accurately. The scores calculated from our previous EWAS conducted in the MCCS ^**12-14**^ may better represent the cumulative lifestyle-related changes to the methylome. Finally, we calculated the age-adjusted measures of biological ageing *PhenoAge* ^**30**^ and *GrimAge*, ^**31**^ based on 513 and 1,030 CpGs, respectively, as done previously.^**32**^ All scores were winsorised at five standard deviations from the mean to minimise the potential influence of outliers, and were then rescaled to Z-scores for better comparability of relative risk estimates.

### Statistical analysis

Correlations between methylation scores were calculated using Spearman correlations in participants selected as controls (to avoid collider bias). The variance explained in each of smoking, BMI and alcohol consumption by their respective methylation scores was also assessed in controls. We used conditional logistic regression to calculate odds ratios, which are estimates of the rate ratios [RRs] when incidence density sampling matching is used, ^**33**^ for the associations between methylation scores, per standard deviation (1SD), and risk of cancer. Two models were used: in <u>Model 1</u>, no covariates were included other than blood cell composition, estimated using the Houseman algorithm;^**34**, **35**^ <u>Model 2</u>, further adjusted for smoking status (never; former; current), smoking pack-years, age at starting (never smoked; age 16 or less; between age 17 and 21; after age 21 years); years since quitting (never smoked; more than 10 years without smoking, between 5 and 10 years without smoking; less than 5 years without smoking), body-mass index (continuous, in kg/m^2^), and alcohol consumption in the past week (continuous, in g/day), a score for physical activity,^**36**^, the Alternate Healthy Eating Index 2010 to reflect overall diet quality,^**37**^ education (score ranging from 1 to 8), socioeconomic status (score ranging from 1 to 10 ^**38**^), and height (continuous, in meters). As a sensitivity analysis, the same models were performed without adjustment for white blood cell composition. These models were used to analyse each cancer separately, and all seven cancer types combined. For the combined analysis, where an individual was diagnosed with several cancers, we included the first diagnosis only (respecting the incidence density sampling procedure), so that participants did not contribute twice to the pooled estimates. Case-control pairs with any missing values for the confounders measured at baseline were excluded and missing values at follow-up (urothelial cancers) were replaced by baseline values; 3% of the initial sample was excluded due to missing values.

We assessed the contribution of methylation scores to the ability of the model to discriminate between cases and controls using the area under the receiver operating characteristic curve (AUC) statistics obtained from unconditional logistic regression models adjusted for the matching variables. We evaluated i) the prediction obtained from models including methylation scores for smoking, BMI, and alcohol consumption instead of the lifestyle variables collected in the cohort; ii) the additional contribution of methylation scores compared with a model with a large collection of traditionally collected health risk factors.

## RESULTS

The characteristics of study participants were described previously ^**32**^ and are shown in **Supplementary Table 2**. The variance explained in the corresponding risk factor was 56% and 28% for Smk-233 and Smk-1061; 15% and 11% for Bmi-1109 and Bmi-85; and 11% and 5% for Alc-450 and Alc-459, respectively (**Table 1)**. The correlations between scores were moderate to high within each lifestyle factor, r=0.62 between Smk-233 and Smk-1061; r=0.46 between Bmi-1109 and Bmi-85, and r=0.27 between Alc-450 and Alc-459. Moderate correlations were also observed across lifestyle factors, in particular between smoking and alcohol scores, between Smk-1,061 and Bmi-85 and between smoking scores and *GrimAge* (**Table 1**). There was little overlap of CpGs between methylation scores, e.g. 17 CpGs between Smk-233 and Smk-1061 **(Supplementary Table 3**).

**Table 2.** Association (rate ratios, 95% confidence intervals) between methylation scores for smoking and cancer risk.

| **Cancer type (N cases)** | **Model 1<sup>a</sup>** |  |  | **Model 2<sup>a</sup>** |  |  |
| --- | --- | --- | --- | --- | --- | --- |
|  | **RR** | **CI95** | **P** | **RR** | **CI95** | **P** |
| **Colorectal (N=814)** |  |  |  |  |  |  |
| **Smk-233** | 1.06 | 0.94-1.19 | 0.32 | 0.91 | 0.75-1.1 | 0.34 |
| **Smk-1061** | 1.06 | 0.94-1.20 | 0.31 | 0.98 | 0.84-1.15 | 0.83 |
| **Gastric (N=166)** |  |  |  |  |  |  |
| **Smk-233** | 1.01 | 0.79-1.29 | 0.95 | 1.10 | 0.73-1.67 | 0.65 |
| **Smk-1061** | 0.95 | 0.72-1.27 | 0.74 | 0.90 | 0.59-1.38 | 0.63 |
| **Kidney (N=139)** |  |  |  |  |  |  |
| **Smk-233** | 1.09 | 0.84-1.42 | 0.50 | 0.82 | 0.46-1.47 | 0.50 |
| **Smk-1061** | 0.97 | 0.73-1.30 | 0.85 | 0.78 | 0.47-1.31 | 0.35 |
| **Lung (N=327)** |  |  |  |  |  |  |
| **Smk-233** | 1.84 | 1.44-2.36 | 2x10<sup>-6</sup> | 1.68 | 1.29-2.19 | 0.0001 |
| **Smk-1061** | 1.59 | 1.26-2.01 | 0.0001 | 1.49 | 1.17-1.90 | 0.001 |
| **MBCN (N=426)** |  |  |  |  |  |  |
| **Smk-233** | 0.97 | 0.80-1.17 | 0.74 | 0.93 | 0.69-1.24 | 0.61 |
| **Smk-1061** | 1.12 | 0.93-1.35 | 0.22 | 1.21 | 0.96-1.52 | 0.11 |
| **Prostate (N=847)** |  |  |  |  |  |  |
| **Smk-233** | 0.89 | 0.79-1.00 | 0.04 | 0.90 | 0.75-1.08 | 0.25 |
| **Smk-1061** | 0.88 | 0.78-1.00 | 0.05 | 0.89 | 0.76-1.04 | 0.15 |
| **Urothelial (N=404)** |  |  |  |  |  |  |
| **Smk-233** | 1.35 | 1.15-1.58 | 0.0002 | 1.16 | 0.9-1.48 | 0.25 |
| **Smk-1061** | 1.47 | 1.24-1.75 | 8x10<sup>-6</sup> | 1.32 | 1.06-1.63 | 0.01 |
| **All types (N=3,000)** |  |  |  |  |  |  |
| **Smk-233** | 1.09 | 1.02-1.15 | 0.006 | 1.06 | 0.97-1.16 | 0.20 |
| **Smk-1061** | 1.11 | 1.04-1.18 | 0.0009 | 1.09 | 1.01-1.18 | 0.02 |

<sup>a</sup> RRs were calculated using conditional logistic regression models. Cases and controls were matched on age, sex, country of birth, sample type and smoking status (for the lung cancer study), Matched pairs were placed consecutively on a same chip of the assay, at random positions.
Model 1: adjusted for white blood cell composition
Model 2: additionally adjusted for smoking status (current/former/never), alcohol consumption (in g/day) and BMI (in kg/m<sup>2</sup>), smoking pack-years, age at starting (never smoked; age 16 or less; between age 17 and 21; after age 21 years); years since quitting (never smoked; more than 10 years without smoking, between 5 and 10 years without smoking; less than 5 years without smoking), a score for physical activity, the alternate healthy eating index to reflect overall diet quality, education (score ranging from 1 to 8), socioeconomic status (score ranging from 1 to 10) and height (continuous, in meters)

**Table 3.** Association (rate ratios, 95% confidence intervals) between methylation scores for BMI and cancer risk.

| **Cancer type (N cases)** | **Model 1<sup>a</sup>** |  |  | **Model 2<sup>a</sup>** |  |  |
| --- | --- | --- | --- | --- | --- | --- |
|  | **RR** | **95% CI** | **P** | **RR** | **95% CI** | **P** |
| **Colorectal (N=814)** |  |  |  |  |  |  |
| **Bmi-1109** | 1.14 | 1.03-1.27 | 0.01 | 1.11 | 0.98-1.26 | 0.10 |
| **Bmi-85** | 1.18 | 1.03-1.35 | 0.01 | 1.11 | 0.96-1.29 | 0.18 |
| **Gastric (N=166)** |  |  |  |  |  |  |
| **Bmi-1109** | 1.27 | 0.98-1.64 | 0.07 | 1.29 | 0.97-1.72 | 0.08 |
| **Bmi-85** | 1.15 | 0.83-1.59 | 0.40 | 1.14 | 0.79-1.63 | 0.49 |
| **Kidney (N=139)** |  |  |  |  |  |  |
| **Bmi-1109** | 1.30 | 1.03-1.63 | 0.03 | 1.22 | 0.92-1.62 | 0.17 |
| **Bmi-85** | 1.16 | 0.82-1.66 | 0.39 | 0.97 | 0.62-1.54 | 0.91 |
| **Lung (N=327)** |  |  |  |  |  |  |
| **Bmi-1109** | 0.90 | 0.76-1.06 | 0.22 | 0.99 | 0.83-1.19 | 0.94 |
| **Bmi-85** | 0.97 | 0.78-1.21 | 0.78 | 1.08 | 0.84-1.38 | 0.57 |
| **MBCN (N=426)** |  |  |  |  |  |  |
| **Bmi-1109** | 0.87 | 0.74-1.01 | 0.07 | 0.81 | 0.68-0.97 | 0.02 |
| **Bmi-85** | 1.25 | 1.02-1.53 | 0.03 | 1.32 | 1.04-1.66 | 0.02 |
| **Prostate (N=847)** |  |  |  |  |  |  |
| **Bmi-1109** | 0.99 | 0.89-1.11 | 0.92 | 0.97 | 0.86-1.10 | 0.61 |
| **Bmi-85** | 0.90 | 0.78-1.05 | 0.18 | 0.90 | 0.77-1.05 | 0.19 |
| **Urothelial (N=404)** |  |  |  |  |  |  |
| **Bmi-1109** | 1.05 | 0.89-1.23 | 0.58 | 1.03 | 0.87-1.23 | 0.72 |
| **Bmi-85** | 1.35 | 1.10-1.66 | 0.004 | 1.23 | 0.98-1.55 | 0.07 |
| **All types (N=3,000)** |  |  |  |  |  |  |
| **Bmi-1109** | 1.03 | 0.98-1.09 | 0.25 | 1.02 | 0.96-1.08 | 0.57 |
| **Bmi-85** | 1.11 | 1.04-1.20 | 0.003 | 1.10 | 1.02-1.19 | 0.01 |

<sup>1</sup> RRs were calculated using conditional logistic regression models. Cases and controls were matched on age, sex, country of birth, sample type and smoking status (for the lung cancer study), Matched pairs were placed consecutively on a same chip of the assay, at random positions.
Model 1: adjusted for white blood cell composition
Model 2: additionally adjusted for smoking status (current/former/never), alcohol consumption (in g/day) and BMI (in kg/m<sup>2</sup>), smoking pack-years, age at starting (never smoked; age 16 or less; between age 17 and 21; after age 21 years); years since quitting (never smoked; more than 10 years without smoking, between 5 and 10 years without smoking; less than 5 years without smoking), a score for physical activity, the alternate healthy eating index to reflect overall diet quality, education (score ranging from 1 to 8), socioeconomic status (score ranging from 1 to 10) and height (continuous, in meters)

The associations of methylation scores for smoking, BMI and alcohol consumption with risk of cancer are shown in **Table 2, 3 and 4**, respectively. The smoking scores showed strong associations with risk of lung and urothelial cancers in unadjusted models (Model 1, adjustment for white blood cell proportions only): lung cancer, Smk-233: RR per SD=1.84, 95%CI: 1.44-2.36, P=2×10^−6^; Smk-1061: RR=1.59, 95%CI: 1.26-2.01, P=0.0001; urothelial cancers: Smk-233: RR=1.35, 95%CI: 1.15-1.58, P=0.0002; Smk-1061: RR=1.47, 95%CI: 1.24-1.75, P=8×10^−6^. There was little attenuation after adjustment for lifestyle factors including several smoking-related variables collected by questionnaires (Model 2), except for Smk33 and urothelial cancer risk (point estimates: lung: Smk-233: 1.68, Smk-1061: 1.49, urothelial: Smk-233: 1.16, Smk-1061: 1.32). Positive but weak associations were observed with risk of cancer overall: unadjusted: Smk-233: 1.09, P=0.006, Smk-1061: 1.11, P=0.0009; adjusted: Smk-233: 1.06, P=0.20, Smk-1061: 1.09, P=0.02. Associations observed without adjustment for cell composition were similar, albeit slightly weaker (**Supplementary Table 4**).

**Table 4.**
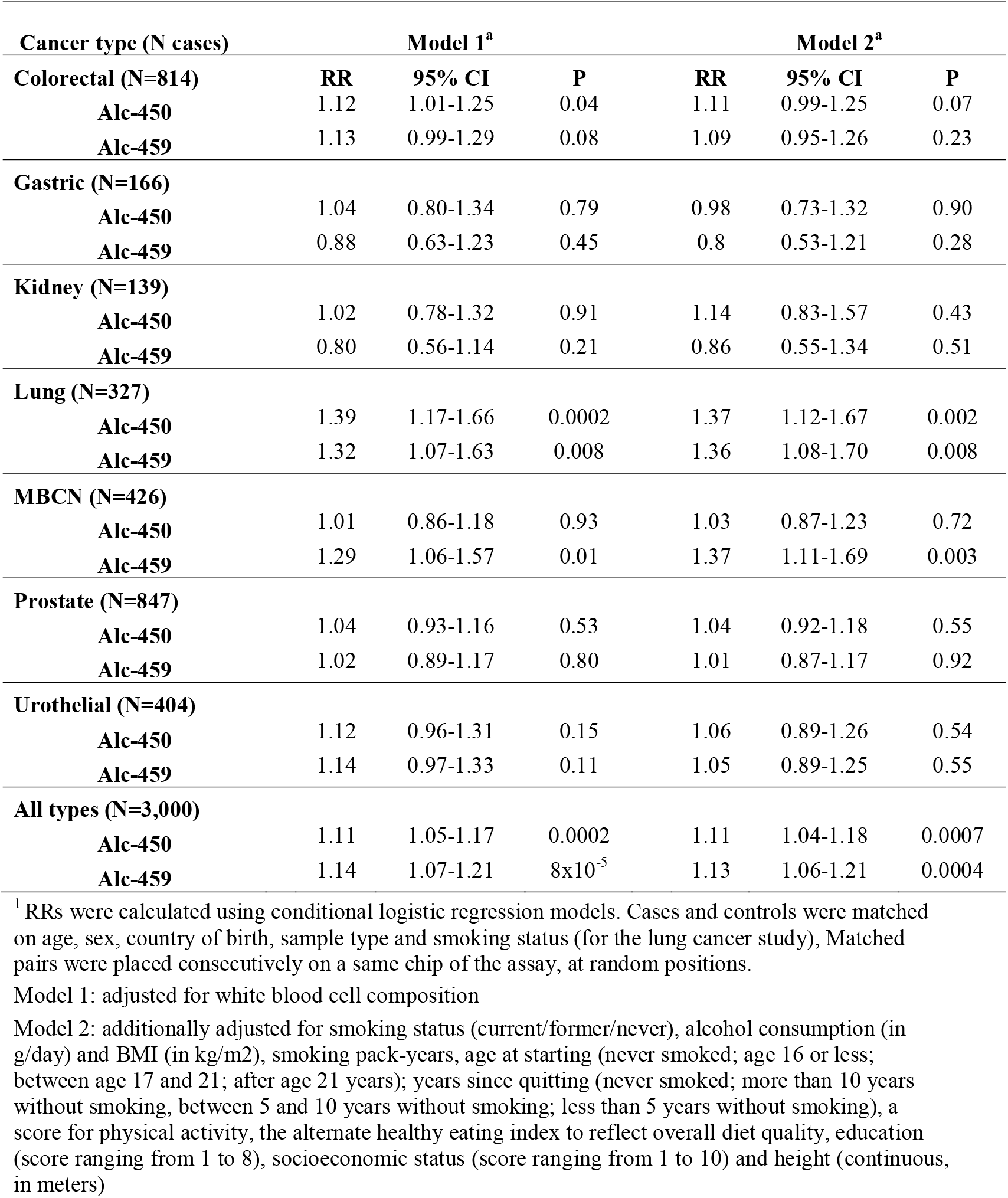
Association (rate ratios, 95% confidence intervals) between methylation scores for alcohol consumption and cancer risk

Methylation scores for BMI generally showed associations with cancer risk in the expected direction but these were weaker than for smoking (**Table 3**). For colorectal cancer, the associations were substantially attenuated in comprehensively adjusted models and were in Model 2: Bmi-1109: RR=1.11, 95%CI: 0.98-1.26, and Bmi-85: RR=1.11, 95%CI: 0.96-1.29. Similar findings were obtained for gastric and kidney cancer risk. Bmi-85 was associated with risk of mature B-cell neoplasms (Model 2: 1.32, 95%CI: 1.04-1.66) and this association was not substantially stronger in models not adjusted for cell proportions, which may substantially confound the associations for this cancer type (RR=1.45, **Supplementary Table 5**). A moderately strong association was observed with risk of urothelial cancer (RR=1.35 and 1.23 in Model 1 and 2, respectively). Bmi-85 showed a stronger association with overall cancer risk than Bmi-1109 (RR=1.10, 95%CI: 1.02-1.19 and RR=1.02, 95%CI: 0.96-1.08, respectively).

**Table 5.**
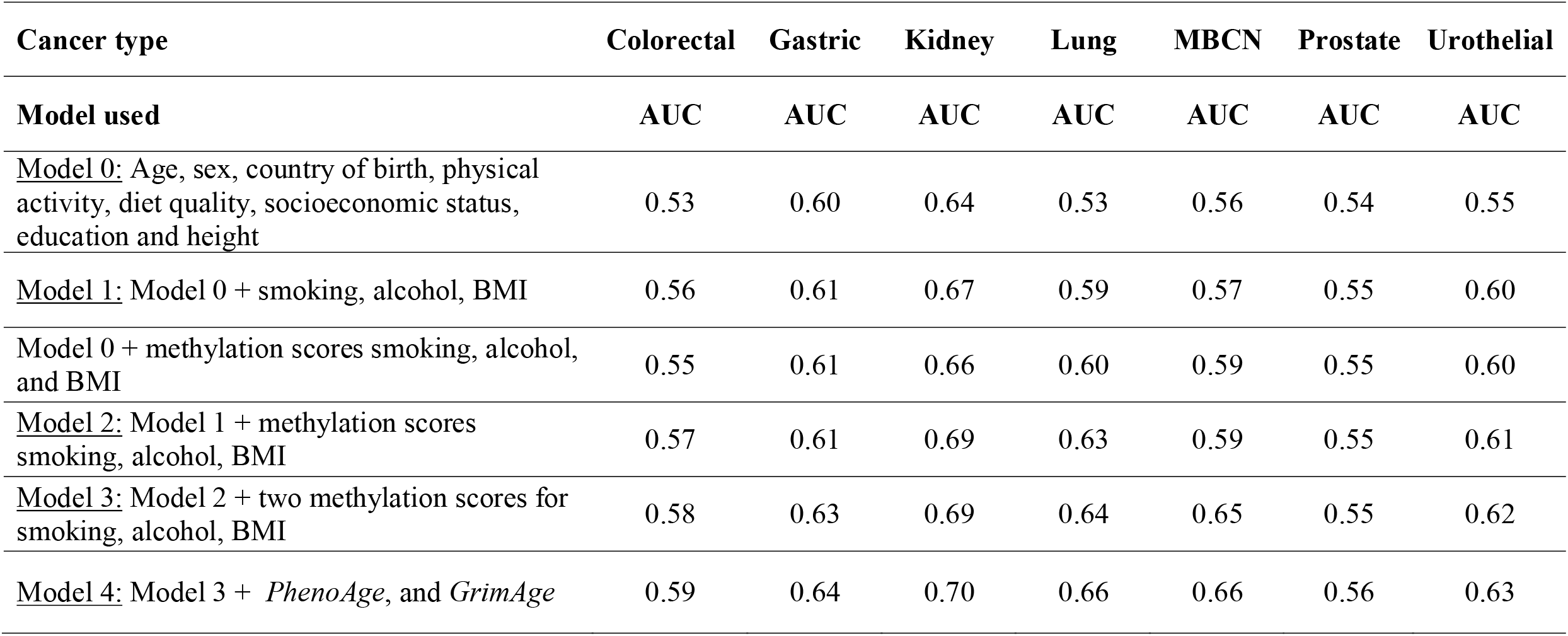
AUCs for various combinations of methylation scores and traditionally-assessed lifestyle variables.

| Cancer type | Colorectal | Gastric | Kidney | Lung | MBCN | Prostate | Urothelial |
| --- | --- | --- | --- | --- | --- | --- | --- |
| Model used | AUC | AUC | AUC | AUC | AUC | AUC | AUC |
| <u>Model 0:</u> Age, sex, country of birth, physical activity, diet quality, socioeconomic status, education and height | 0.53 | 0.60 | 0.64 | 0.53 | 0.56 | 0.54 | 0.55 |
| <u>Model 1:</u> Model 0 + smoking, alcohol, BMI | 0.56 | 0.61 | 0.67 | 0.59 | 0.57 | 0.55 | 0.60 |
| Model 0 + methylation scores smoking, alcohol, and BMI | 0.55 | 0.61 | 0.66 | 0.60 | 0.59 | 0.55 | 0.60 |
| <u>Model 2:</u> Model 1 + methylation scores smoking, alcohol, BMI | 0.57 | 0.61 | 0.69 | 0.63 | 0.59 | 0.55 | 0.61 |
| <u>Model 3:</u> Model 2 + two methylation scores for smoking, alcohol, BMI | 0.58 | 0.63 | 0.69 | 0.64 | 0.65 | 0.55 | 0.62 |
| <u>Model 4:</u> Model 3 + *PhenoAge*, and *GrimAge* | 0.59 | 0.64 | 0.70 | 0.66 | 0.66 | 0.56 | 0.63 |

For alcohol consumption, we found that all scores were associated with risk of lung cancer (Model 2: RR=1.37, P=0.002 and RR=1.36, P=0.008 for Alc-450 and Alc-459, respectively), **Table 4**. Small associations were observed with risk of colorectal cancer (RR=1.11 and RR=1.09) and a relatively large association for Alc-459 with risk of B-cell lymphoma RR=1.37, 95%CI: 1.11-1.69. For cancer overall, the risk was increased by 13% (4% to 18%) and 11% (6% to 21%) per SD increase in Alc-450 and Alc-459, respectively, after adjustment for cohort-collected sociodemographic, anthropometric and lifestyle variables; these associations were virtually the same as in unadjusted models, without adjustment for cell proportions, or using the scores of Liu et al. (**Supplementary Tables 6 and 7)**. For the scores of Liu et al., the associations did not appear larger for the scores including more CpGs, for example for cancer overall: Alc-144: RR=1.08, 95%CI: 1.02-1.15; Alc-78: RR=1.12, 95%CI: 1.06-1.19; Alc-23: RR=1.15, 95%CI: 1.08-1.22 and Alc-5: RR=1.14, 95%CI: 1.07-1.21 (**Supplementary Table 7**).

Results for the contribution of methylation scores to the prediction of cancer risk are shown in **Table 5**. The AUCs of models with methylation scores instead of traditionally assessed smoking, BMI, and alcohol consumption appeared similar for all cancer types and were somewhat higher for lung cancer (0.60 vs 0.59) and mature B-cell neoplasms (0.59 vs 0.57). The addition of methylation scores to variables that were collected via questionnaires or measured in the MCCS nevertheless resulted in relatively small AUC gains: models including two methylation scores for each of smoking, BMI, and alcohol consumption, *GrimAge* and *PhenoAge* compared with models without: colorectal cancer: AUC=0.59 vs 0.56, gastric: 0.64 vs 0.61, kidney: 0.70 vs 0.67, lung: 0.66 vs 0.59, mature B-cell neoplasms: 0.66 vs 0.57, prostate: 0.56 vs 0.55, urothelial: 0.63 vs 0.60.

## DISCUSSION

We calculated various methylation scores for smoking, alcohol consumption and BMI, the main lifestyle factors that cause changes to DNA methylation and increase the risk of cancer.^**12-14**, **39-41**^ Several of these were associated with certain types of cancer and associations were only moderately attenuated after adjustment for sociodemographic, anthropometric and lifestyle factors, including several questionnaire-collected variables relating to smoking history (pack-years, age at starting, time since quitting). These associations were nevertheless generally relatively small, except for smoking scores and lung and urothelial cancer risk, for which we reported some of the findings previously.^**42**^ Reasonably large associations were also observed for methylation scores for BMI and alcohol consumption with risk of mature B-cell lymphoma (RR per SD∼1.3-1.4), but these should be interpreted with some caution given the strongly modified blood methylation profiles found in B-cell neoplasms, ^**26**^ and the fact that lifestyle is thought to only moderately increase risk for these types of cancer.^**43**^ For risk of cancer overall, the methylation scores based on weights derived from EWAS conducted in the MCCS ^**12-14**^ produced somewhat stronger associations with cancer risk than those obtained via regularised regression,^**19**^ being estimated in comprehensively adjusted models at 9%, 10%, and 13% increase, compared with 6%, 2% and 11%, per SD of smoking, BMI, and alcohol methylation scores, respectively. That associations appeared stronger with these scores than with those of McCartney et al. ^**19**^ may be explained in part by the way the scores were built (i.e. ‘cumulative damage’ provided by the weighted average for all CpGs identified as top-ranked associations between health risk factors and DNA methylation, instead of ‘best predictor’, obtained via regularised regression, which aims to maximise prediction), and perhaps more importantly because methylation at all CpGs they included was found to be strongly associated with these lifestyle factors in the samples used in this study.^**12-14**^. Consistent with this, the predictor found by Liu et al. ^**11**^ to be the best predictor of alcohol consumption in external data (based on 144 CpGs) performed less well than the reduced versions of it in terms of strength of association with cancer risk. In line with the relatively small adjusted associations we observed, these scores, in addition to *PhenoAge* ^**30**, **32**^ and *GrimAge*,^**31**, **32**^ only provided moderate added discrimination between cases and controls (∼3% for colorectal, gastric, kidney and urothelial cancers, ∼7% for lung cancer and ∼8% for mature B-cell lymphomas).

The two main strengths of this study are i) its prospective design using incidence density sampling with improved control for confounding and efficiency by carefully matching cases and controls on several cancer-associated factors, including smoking history for the lung cancer study, as well as very good control for batch effects provided by placing matched pairs on the same slide of the array; and ii) its large sample size (3,123 cancer cases) and the inclusion of several cancer types. The main limitation of our study is that it does not provide a ‘single best’ or integrated / synthetic methylation score representative of each lifestyle factor. It could also be that while methylation scores provided a good summary of the effect of lifestyle on the methylome, they were not able to capture the methylation patterns specific to risk for each cancer, as it is likely that methylation changes in certain genomic regions may have different implications regarding susceptibility to different cancers.

To our knowledge, our study is novel in the field of cancer, and reports of similar attempts to assess other health outcomes are limited. In the study by McCartney et al.,^**19**^ the authors examined all-cause mortality and showed it was associated with their BMI methylation score (HR=1.23, 95%CI: 1.06-1.42, after adjustment for age, sex, white blood cell proportions, smoking methylation score, BMI polygenic score and measured BMI). Somewhat weaker associations were observed for waist-to-hip ratio and body fat percentage, which we did not include in our study. A negative association was observed for methylation-predicted total cholesterol. The association for their alcohol consumption methylation score was similar to those observed in our study (HR=1.11, 95%CI: 0.97-1.28). A strong association was observed for the smoking methylation score, in particular after adjustment for BMI methylation score (HR=1.57, 95%CI: 1.39-1.78). Other studies published by the same group showed increased prediction of several health-related outcomes for their BMI methylation score ^**44**^ and their smoking methylation score ^**45**^. In the study by Langdon et al., ^**46**^ methylation scores were evaluated in participants with oropharyngeal cancer and moderately large associations (HR∼1.2-1.3) with survival were observed for some scores in adjusted models, with modest improvements in terms of prediction. Three other scores were evaluated in that study ^**10**, **47**, **48**^ as well as just methylation at cg05575921 (*AHRR*, smoking), ^**12**^ for which the results were similar to those we considered. Other attempts were also made to use individual, or a restricted set of, smoking-associated methylation marks to predict lung cancer or other smoking-related outcomes, focusing on the strongest observed associations (e.g. *AHRR*), for example in the studies by Baglietto et al. including MCCS data, ^**23**^ Zhang et al., ^**49**, **50**^ and others. ^**51-54**^

We conclude that methylation scores for smoking, BMI, and alcohol consumption show some associations with risk of cancer, independently of many health-related variables, but these were generally relatively small. While these scores could potentially replace the variables routinely collected in cohorts, their added contribution to the prediction of cancer is modest.

## Data Availability

The data and code used in this study are available from the corresponding author upon reasonable request.

## Acknowledgements

Cases were ascertained through the Victorian Cancer Registry (VCR) and the Australian Cancer Database (Australian Institute of Health and Welfare).

## Data Availability Statement

The methylation array data that support the findings of this study will be deposited to an appropriate (restricted-access) public repository for public release upon publication.

## Ethics Statement

Participants provided informed consent in accordance with the Declaration of Helsinki and the study was approved by Cancer Council Victoria’s Human Research Ethics Committee.

## Abbreviations

AHRR: aryl hydrocarbon receptor repressor
AUC: area under the receiver operating characteristic curve
BMI: body mass index
EWAS: epigenome-wide association study
MCCS: Melbourne Collaborative Cohort Study
MS: methylation score
SD: standard deviation,

