## Supplementary Methods and Tables for "Methylation scores for smoking, alcohol consumption, and body mass index and risk of seven types of cancer"

**DNA extraction and bisulfite conversion, and DNA methylation data processing**

*Bisulphite conversion of DNA and Illumina Infinium methylation assay*

Genomic DNA was extracted from mononuclear cells using the QIAamp 96 DNA blood kit (Qiagen), and from Guthrie card samples as previously described **^1^****.** Briefly, twenty blood spots of 3.2 mm diameter were punched from the Guthrie card and lysed in phosphate buffered saline using TissueLyser (Qiagen). The resulting supernatant was processed using Qiagen mini spin columns according to the manufacturer’s protocol. The quality and quantity of DNA was assessed using the Quant-iT™ Picogreen® dsDNA assay measured on the Qubit® Fluorometer (Life Technologies, Grand Island, NY), with a minimum of 0.3 μg DNA considered acceptable for methylation analysis. Genomic DNA was bisulphite converted using the EZ DNA Methylation-Gold kit (Zymo Research, Irvine, CA) following the manufacturer’s instructions. The Illumina Infinium HumanMethylation450K BeadChip (HM450K) array (Illumina, Inc.; San Diego, CA, USA) was used to measure DNA methylation. This array covers 99% of the RefSeq genes and uses a combination of two distinct probe types (Infinium I and II) to detect the methylation status of 485,577 CpGs in the human genome at a single base resolution **^2^****.** Samples in each nested case-control study and the longitudinal study were assayed at non-overlapping time periods. In each study, samples were randomly assigned to chips and processed as per the Illumina protocol. All laboratory work, including DNA extraction, bisulphite conversion and Infinium assaying was performed at the Genetic Epidemiology Laboratory, University of Melbourne.

***Normalization and quality control of methylation data***

The same normalization and quality control procedures were applied to methylation data in all eight studies (case-control and longitudinal). Raw .IDAT files were imported into R using the *minfi* package **^3^****.** Illumina’s background correction was applied based on internal control probes. Subset-quantile within-array normalisation (SWAN) was used to correct for the technical variability between the two probe types of the HM450K array **^4^**. The ‘getSex’ function of the *minfi* package **^3^** was used to predict sex for each sample. Samples for which predicted sex was inconsistent with recorded sex were excluded from further analyses. For each sample, each CpG with a detection *P* value >0.01 was assigned as missing. Samples with missing values for more than 5% of probes were excluded. CpG sites were excluded if they were missing for more than 20% of samples. β-values, which range from zero to one and correspond to the percentage of methylation, were calculated for each CpG using *minfi*. β-values were transformed into M-values using the formula: $M= {log}_{2}(\frac{\beta}{1-\beta})$. **^5^**.

**Supplementary Table 1.** Median and interquartile range of time to diagnosis by cancer type

| **Cancer type** | **Median time-to-diagnosis** | **Interquartile range** |
| --- | --- | --- |
| Colorectal cancer | 9.3 | 5.4-13.2 |
| Gastric cancer | 11.2 | 6.5-16.2 |
| Kidney cancer | 11.2 | 5.7-14.2 |
| Lung cancer | 10.1 | 5.7-13.4 |
| Mature B-cell neoplasms | 10.6 | 6.2-14.3 |
| Prostate cancer | 10.5 | 6.2-14.0 |
| Urothelial cancer | 6.3 | 3.7-10.7 |
| Overall | 9.4 | 5.2-13.3 |

**Supplementary Table 2.** Characteristics of the study sample, seven case-control studies nested within the Melbourne Collaborative Cohort Study (N=41,513)

|  | **Controls** | **Cases** |
| --- | --- | --- |
| **Cancer type (N)** |  |  |
| Colorectal cancer | 813 | 813 |
| Gastric cancer | 165 | 165 |
| Kidney cancer | 139 | 139 |
| Lung cancer | 327 | 327 |
| Mature B-cell neoplasms | 423 | 423 |
| Prostate cancer | 846 | 846 |
| Urothelial cancers | 404 | 404 |
| **Matching variables** |  |  |
| Age at blood draw (years), median [IQR] | 61 [54-66] | 61 [54-66] |
| Sex: male, N (%) | 2,163 (61%) | 2,163 (61%) |
| Sex: female, N (%) | 1,366 (39%) | 1,366 (39%) |
| Country of birth: Australia/NZ, N (%) | 2,400 (68%) | 2,416 (68%) |
| Northern Europe, N (%) | 237 (7%) | 230 (7%) |
| Southern Europe, N (%) | 892 (25%) | 883 (25%) |
| Blood sample type: Dried blood spots, N (%) | 2,463 (70%) | 2,463 (70%) |
| Peripheral blood mononuclear cells, N (%) | 877 (25%) | 877 (25%) |
| Buffy coats, N (%) | 189 (5%) | 189 (5%) |
| **Potential confounders** |  |  |
| Smoking: current, N (%) | 485 (14%) | 406 (15%) |
| Smoking: former, N (%) | 1,345 (38%) | 1,392 (39%) |
| Smoking: never, N (%) | 1,699 (48%) | 1,625 (46%) |
| Smoking pack-years, median [IQR] | 11 [0-500] | 39 [0-580] |
| Age at starting: never smoked, N (%) | 1,699 (48%) | 1,625 (46%) |
| Age at starting: ≤16 years, N (%) | 682 (19%) | 697 (20%) |
| Age at starting: 17-21 years, N (%) | 815 (23%) | 899 (25%) |
| Age at starting: ≥22 years, N (%) | 333 (9%) | 308 (9%) |
| Time since quitting: never smoked, N (%) | 1,699 (48%) | 1,625 (46%) |
| Time since quitting: <5 years, N (%) | 672 (19%) | 725 (21%) |
| Time since quitting: 5-10 years, N (%) | 932 (26%) | 938 (27%) |
| Time since quitting: >10 years, N (%) | 226 (6%) | 241 (7%) |
| Body mass index (kg/m^2^), median [IQR] | 27 [24-29] | 27 [24-30] |
| Height (m), median [IQR] | 167 [160-174] | 167 [161-174] |
| Alcohol consumption (g/day), median [IQR] | 4 [0-17] | 4 [0-18] |
| Diet quality: AHEI-2010, median [IQR] | 64 [56-72] | 64 [56-73] |
| Physical activity score, median [IQR] | 2 [1-4] | 2 [1-4] |
| Education score, median [IQR] | 4 [4-6] | 4 [4-6] |
| Socioeconomic status: SEIFA-10, median [IQR] | 5 [3-8] | 6 [3-9] |

**Supplementary Table 3.** Number of overlapping CpG sites between methylation scores

|  | **Smk-233** | **Smk-1061** | **Bmi-1109** | **Bmi-85** | **Alc-450** | **Alc-459** | **Alc-144** | **Alc-78** | **Alc-23** | **Alc-5** | ***PhenoAge*** | ***GrimAge*** |
| --- | --- | --- | --- | --- | --- | --- | --- | --- | --- | --- | --- | --- |
| **Smk-233** |  | 17 | 11 | 0 | 4 | 3 | 0 | 0 | 0 | 0 | 1 | 8 |
| **Smk-1061** |  |  | 35 | 10 | 8 | 69 | 9 | 5 | 0 | 0 | 5 | 36 |
| **Bmi-1109** |  |  |  | 25 | 12 | 14 | 7 | 3 | 2 | 1 | 3 | 26 |
| **Bmi-85** |  |  |  |  | 5 | 12 | 5 | 3 | 2 | 1 | 1 | 12 |
| **Alc-450** |  |  |  |  |  | 16 | 6 | 5 | 4 | 3 | 1 | 5 |
| **Alc-459** |  |  |  |  |  |  | 42 | 29 | 8 | 4 | 1 | 13 |
| **Alc-144** |  |  |  |  |  |  |  | 78 | 23 | 5 | 1 | 2 |
| **Alc-78** |  |  |  |  |  |  |  |  | 23 | 5 | 1 | 2 |
| **Alc-23** |  |  |  |  |  |  |  |  |  | 5 | 1 | 2 |
| **Alc-5** |  |  |  |  |  |  |  |  |  |  | 1 | 1 |
| ***PhenoAge^a^*** |  |  |  |  |  |  |  |  |  |  |  | 8 |
| ***GrimAge^b^*** |  |  |  |  |  |  |  |  |  |  |  |  |

^a^based on 513 CpGs,

^b^ based on 1,030 CpGs

**Supplementary Table 4**. Association (rate ratios, 95% confidence intervals) between methylation scores for smoking and cancer risk, without adjustment for white blood cell proportions

| **Cancer type** | **Model 1^a^** | | | |  | **Model 2^a^** | | |
| --- | --- | --- | --- | --- | --- | --- | --- | --- |
| **COLORECTAL** |  | **RR** | **CI95** | **P** |  | **RR** | **CI95** | **P** |
| **Smk-233** |  | 1.05 | 0.93-1.17 | 0.42 |  | 0.89 | 0.74-1.07 | 0.23 |
| **Smk-1061** |  | 1.02 | 0.92-1.14 | 0.69 |  | 0.94 | 0.83-1.08 | 0.40 |
| **GASTRIC** |  |  |  |  |  |  |  |  |
| **Smk-233** |  | 1.04 | 0.82-1.31 | 0.77 |  | 1.11 | 0.74-1.67 | 0.60 |
| **Smk-1061** |  | 0.85 | 0.66-1.10 | 0.22 |  | 0.77 | 0.54-1.10 | 0.15 |
| **KIDNEY** |  |  |  |  |  |  |  |  |
| **Smk-233** |  | 1.06 | 0.82-1.36 | 0.66 |  | 0.83 | 0.47-1.44 | 0.50 |
| **Smk-1061** |  | 0.93 | 0.71-1.21 | 0.57 |  | 0.79 | 0.51-1.22 | 0.28 |
| **LUNG** |  |  |  |  |  |  |  |  |
| **Smk-233** |  | 1.84 | 1.44-2.35 | 1x10^-6^ |  | 1.66 | 1.28-2.15 | 0.0001 |
| **Smk-1061** |  | 1.44 | 1.16-1.78 | 0.0008 |  | 1.38 | 1.10-1.72 | 0.005 |
| **MBCN** |  |  |  |  |  |  |  |  |
| **Smk-233** |  | 0.91 | 0.76-1.10 | 0.33 |  | 0.86 | 0.66-1.12 | 0.25 |
| **Smk-1061** |  | 1.32 | 1.13-1.55 | 0.0004 |  | 1.48 | 1.23-1.78 | 3x10^-5^ |
| **PROSTATE** |  |  |  |  |  |  |  |  |
| **Smk-233** |  | 0.91 | 0.81-1.01 | 0.08 |  | 0.92 | 0.76-1.10 | 0.34 |
| **Smk-1061** |  | 0.90 | 0.80-1.01 | 0.06 |  | 0.91 | 0.79-1.05 | 0.19 |
| **UROTHELIAL** |  |  |  |  |  |  |  |  |
| **Smk-233** |  | 1.36 | 1.17-1.58 | 7x10^-5^ |  | 1.16 | 0.91-1.48 | 0.24 |
| **Smk-1061** |  | 1.39 | 1.18-1.63 | 6x10^-5^ |  | 1.21 | 0.99-1.47 | 0.06 |
| **ALL CANCERS** |  |  |  |  |  |  |  |  |
| **Smk-233** |  | 1.07 | 1.01-1.14 | 0.01 |  | 1.04 | 0.96-1.14 | 0.35 |
| **Smk-1061** |  | 1.09 | 1.03-1.15 | 0.004 |  | 1.07 | 1.00-1.14 | 0.05 |

^a^ RRs were calculated using conditional logistic regression models. Cases and controls were matched on age, sex, country of birth, sample type and smoking status (for the lung cancer study), Matched pairs were placed consecutively on a same chip of the assay, at random positions.

Model 1: no adjustment

Model 2: additionally adjusted for smoking status (current/former/never), alcohol consumption (in g/day) and BMI (in kg/m2), smoking pack-years, age at starting (never smoked; age 16 or less; between age 17 and 21; after age 21 years); years since quitting (never smoked; more than 10 years without smoking, between 5 and 10 years without smoking; less than 5 years without smoking), a score for physical activity, the alternate healthy eating index to reflect overall diet quality, education (score ranging from 1 to 8), socioeconomic status (score ranging from 1 to 10) and height (continuous, in meters)

**Supplementary Table 5**. Association (rate ratios, 95% confidence intervals) between methylation scores for BMI and cancer risk, without adjustment for white blood cell proportions

| **Cancer type** | **Model 1^a^** | | | |  | **Model 2^a^** | | |
| --- | --- | --- | --- | --- | --- | --- | --- | --- |
| **COLORECTAL** |  | **RR** | **95% CI** | **P** |  | **RR** | **95% CI** | **P** |
| **Bmi-1109** |  | 1.12 | 1.01-1.25 | 0.03 |  | 1.08 | 0.96-1.22 | 0.20 |
| **Bmi-85** |  | 1.10 | 0.99-1.22 | 0.09 |  | 1.05 | 0.94-1.18 | 0.40 |
| **GASTRIC** |  |  |  |  |  |  |  |  |
| **Bmi-1109** |  | 1.26 | 0.98-1.61 | 0.07 |  | 1.30 | 0.98-1.72 | 0.07 |
| **Bmi-85** |  | 0.90 | 0.70-1.14 | 0.38 |  | 0.91 | 0.69-1.19 | 0.49 |
| **KIDNEY** |  |  |  |  |  |  |  |  |
| **Bmi-1109** |  | 1.29 | 1.03-1.61 | 0.03 |  | 1.20 | 0.91-1.57 | 0.19 |
| **Bmi-85** |  | 1.13 | 0.85-1.50 | 0.41 |  | 0.99 | 0.69-1.42 | 0.95 |
| **LUNG** |  |  |  |  |  |  |  |  |
| **Bmi-1109** |  | 0.90 | 0.77-1.06 | 0.21 |  | 0.98 | 0.82-1.17 | 0.86 |
| **Bmi-85** |  | 0.95 | 0.80-1.13 | 0.58 |  | 1.04 | 0.86-1.25 | 0.72 |
| **MBCN** |  |  |  |  |  |  |  |  |
| **Bmi-1109** |  | 0.82 | 0.71-0.95 | 0.008 |  | 0.79 | 0.67-0.93 | 0.005 |
| **Bmi-85** |  | 1.33 | 1.13-1.57 | 6x10^-4^ |  | 1.45 | 1.20-1.74 | 7x10^-5^ |
| **PROSTATE** |  |  |  |  |  |  |  |  |
| **Bmi-1109** |  | 0.99 | 0.89-1.11 | 0.88 |  | 0.97 | 0.85-1.09 | 0.57 |
| **Bmi-85** |  | 0.94 | 0.84-1.05 | 0.26 |  | 0.95 | 0.84-1.07 | 0.37 |
| **UROTHELIAL** |  |  |  |  |  |  |  |  |
| **Bmi-1109** |  | 1.05 | 0.89-1.23 | 0.57 |  | 1.03 | 0.87-1.22 | 0.76 |
| **Bmi-85** |  | 1.13 | 0.97-1.32 | 0.11 |  | 1.07 | 0.9-1.26 | 0.45 |
| **ALL CANCERS** |  |  |  |  |  |  |  |  |
| **Bmi-1109** |  | 1.02 | 0.97-1.08 | 0.42 |  | 1.01 | 0.95-1.07 | 0.79 |
| **Bmi-85** |  | 1.06 | 1.00-1.12 | 0.05 |  | 1.05 | 0.99-1.12 | 0.08 |

^1^ RRs were calculated using conditional logistic regression models. Cases and controls were matched on age, sex, country of birth, sample type and smoking status (for the lung cancer study), Matched pairs were placed consecutively on a same chip of the assay, at random positions.

Model 1: no adjustment

Model 2: additionally adjusted for smoking status (current/former/never), alcohol consumption (in g/day) and BMI (in kg/m2), smoking pack-years, age at starting (never smoked; age 16 or less; between age 17 and 21; after age 21 years); years since quitting (never smoked; more than 10 years without smoking, between 5 and 10 years without smoking; less than 5 years without smoking), a score for physical activity, the alternate healthy eating index to reflect overall diet quality, education (score ranging from 1 to 8), socioeconomic status (score ranging from 1 to 10) and height (continuous, in meters)

**Supplementary Table 6**. Association (rate ratios, 95% confidence intervals) between methylation scores for alcohol consumption and cancer risk, without adjustment for white blood cell proportions

| **Cancer type** | **Model 1^a^** | | | |  | **Model 2^a^** | | |
| --- | --- | --- | --- | --- | --- | --- | --- | --- |
| **COLORECTAL** |  | **RR** | **95% CI** | **P** |  | **RR** | **95% CI** | **P** |
| **Alc-450** |  | 1.12 | 1.01-1.25 | 0.03 |  | 1.12 | 1.00-1.25 | 0.05 |
| **Alc-459** |  | 1.18 | 1.05-1.32 | 0.006 |  | 1.16 | 1.03-1.31 | 0.01 |
| **GASTRIC** |  |  |  |  |  |  |  |  |
| **Alc-450** |  | 1.06 | 0.83-1.36 | 0.61 |  | 1.03 | 0.78-1.35 | 0.85 |
| **Alc-459** |  | 0.92 | 0.70-1.22 | 0.57 |  | 0.88 | 0.63-1.22 | 0.44 |
| **KIDNEY** |  |  |  |  |  |  |  |  |
| **Alc-450** |  | 1.06 | 0.82-1.36 | 0.66 |  | 1.17 | 0.87-1.59 | 0.29 |
| **Alc-459** |  | 0.92 | 0.69-1.23 | 0.58 |  | 0.97 | 0.68-1.38 | 0.87 |
| **LUNG** |  |  |  |  |  |  |  |  |
| **Alc-450** |  | 1.34 | 1.13-1.58 | 0.0007 |  | 1.33 | 1.10-1.61 | 0.004 |
| **Alc-459** |  | 1.27 | 1.06-1.52 | 0.01 |  | 1.29 | 1.05-1.57 | 0.01 |
| **MBCN** |  |  |  |  |  |  |  |  |
| **Alc-450** |  | 0.92 | 0.79-1.06 | 0.26 |  | 0.94 | 0.80-1.11 | 0.47 |
| **Alc-459** |  | 1.35 | 1.15-1.58 | 0.0002 |  | 1.42 | 1.20-1.67 | 5x10^-5^ |
| **PROSTATE** |  |  |  |  |  |  |  |  |
| **Alc-450** |  | 1.05 | 0.94-1.17 | 0.35 |  | 1.05 | 0.93-1.19 | 0.41 |
| **Alc-459** |  | 1.04 | 0.92-1.18 | 0.50 |  | 1.02 | 0.90-1.17 | 0.72 |
| **UROTHELIAL** |  |  |  |  |  |  |  |  |
| **Alc-450** |  | 1.13 | 0.97-1.31 | 0.13 |  | 1.06 | 0.90-1.26 | 0.49 |
| **Alc-459** |  | 1.14 | 0.99-1.31 | 0.08 |  | 1.07 | 0.92-1.24 | 0.41 |
| **ALL CANCERS** |  |  |  |  |  |  |  |  |
| **Alc-450** |  | 1.09 | 1.03-1.15 | 0.002 |  | 1.09 | 1.03-1.16 | 0.004 |
| **Alc-459** |  | 1.15 | 1.08-1.22 | 3x10^-6^ |  | 1.14 | 1.08-1.22 | 1x10^-5^ |

^1^ RRs were calculated using conditional logistic regression models. Cases and controls were matched on age, sex, country of birth, sample type and smoking status (for the lung cancer study), Matched pairs were placed consecutively on a same chip of the assay, at random positions.

Model 1: no adjustment

Model 2: additionally adjusted for smoking status (current/former/never), alcohol consumption (in g/day) and BMI (in kg/m2), smoking pack-years, age at starting (never smoked; age 16 or less; between age 17 and 21; after age 21 years); years since quitting (never smoked; more than 10 years without smoking, between 5 and 10 years without smoking; less than 5 years without smoking), a score for physical activity, the alternate healthy eating index to reflect overall diet quality, education (score ranging from 1 to 8), socioeconomic status (score ranging from 1 to 10) and height (continuous, in meters)

**Supplementary Table 7**. Association (rate ratios, 95% confidence intervals) between methylation scores for alcohol consumption (scores by Liu et al.) and cancer risk

| **Cancer type** | **Model 1^a^** | | | |  | **Model 2^a^** | | |
| --- | --- | --- | --- | --- | --- | --- | --- | --- |
| **COLORECTAL** |  | **RR** | **95% CI** | **P** |  | **RR** | **95% CI** | **P** |
| **Alc-144** |  | 1.18 | 1.04-1.33 | 0.008 |  | 1.16 | 1.02-1.32 | 0.03 |
| **Alc-78** |  | 1.14 | 1.02-1.29 | 0.03 |  | 1.11 | 0.97-1.26 | 0.12 |
| **Alc-23** |  | 1.12 | 1.00-1.25 | 0.06 |  | 1.08 | 0.95-1.22 | 0.25 |
| **Alc-5** |  | 1.10 | 0.98-1.23 | 0.10 |  | 1.06 | 0.94-1.19 | 0.37 |
| **GASTRIC** |  |  |  |  |  |  |  |  |
| **Alc-144** |  | 1.07 | 0.83-1.38 | 0.58 |  | 0.97 | 0.72-1.30 | 0.83 |
| **Alc-78** |  | 1.13 | 0.88-1.46 | 0.34 |  | 1.05 | 0.78-1.42 | 0.75 |
| **Alc-23** |  | 1.18 | 0.91-1.53 | 0.21 |  | 1.16 | 0.85-1.57 | 0.35 |
| **Alc-5** |  | 1.18 | 0.91-1.53 | 0.20 |  | 1.17 | 0.86-1.60 | 0.31 |
| **KIDNEY** |  |  |  |  |  |  |  |  |
| **Alc-144** |  | 0.88 | 0.66-1.17 | 0.37 |  | 0.96 | 0.69-1.34 | 0.81 |
| **Alc-78** |  | 0.98 | 0.74-1.31 | 0.90 |  | 1.10 | 0.77-1.58 | 0.59 |
| **Alc-23** |  | 1.02 | 0.75-1.39 | 0.89 |  | 1.13 | 0.77-1.67 | 0.53 |
| **Alc-5** |  | 1.06 | 0.78-1.45 | 0.69 |  | 1.17 | 0.80-1.73 | 0.42 |
| **LUNG** |  |  |  |  |  |  |  |  |
| **Alc-144** |  | 1.25 | 1.06-1.49 | 0.01 |  | 1.3 | 1.07-1.58 | 0.009 |
| **Alc-78** |  | 1.31 | 1.10-1.55 | 0.002 |  | 1.34 | 1.11-1.63 | 0.002 |
| **Alc-23** |  | 1.28 | 1.08-1.52 | 0.005 |  | 1.28 | 1.07-1.54 | 0.008 |
| **Alc-5** |  | 1.22 | 1.03-1.44 | 0.02 |  | 1.23 | 1.02-1.48 | 0.03 |
| **MBCN** |  |  |  |  |  |  |  |  |
| **Alc-144** |  | 1.12 | 0.94-1.33 | 0.20 |  | 1.14 | 0.95-1.37 | 0.16 |
| **Alc-78** |  | 1.16 | 0.98-1.38 | 0.09 |  | 1.20 | 1.00-1.44 | 0.06 |
| **Alc-23** |  | 1.27 | 1.06-1.52 | 0.01 |  | 1.35 | 1.12-1.64 | 0.002 |
| **Alc-5** |  | 1.29 | 1.07-1.55 | 0.006 |  | 1.40 | 1.15-1.71 | 0.0009 |
| **PROSTATE** |  |  |  |  |  |  |  |  |
| **Alc-144** |  | 0.94 | 0.83-1.05 | 0.27 |  | 0.91 | 0.8-1.04 | 0.18 |
| **Alc-78** |  | 0.99 | 0.88-1.11 | 0.87 |  | 0.98 | 0.86-1.11 | 0.72 |
| **Alc-23** |  | 1.04 | 0.93-1.17 | 0.44 |  | 1.04 | 0.92-1.18 | 0.53 |
| **Alc-5** |  | 1.05 | 0.94-1.17 | 0.42 |  | 1.04 | 0.92-1.18 | 0.49 |
| **UROTHELIAL** |  |  |  |  |  |  |  |  |
| **Alc-144** |  | 1.06 | 0.91-1.24 | 0.48 |  | 1.03 | 0.87-1.22 | 0.73 |
| **Alc-78** |  | 1.12 | 0.97-1.31 | 0.13 |  | 1.10 | 0.93-1.30 | 0.26 |
| **Alc-23** |  | 1.16 | 1.00-1.35 | 0.06 |  | 1.13 | 0.96-1.33 | 0.13 |
| **Alc-5** |  | 1.17 | 1.00-1.35 | 0.05 |  | 1.14 | 0.97-1.34 | 0.12 |
| **ALL CANCERS** |  |  |  |  |  |  |  |  |
| **Alc-144** |  | 1.08 | 1.02-1.15 | 0.006 |  | 1.08 | 1.02-1.15 | 0.01 |
| **Alc-78** |  | 1.12 | 1.06-1.19 | 6x10^-5^ |  | 1.12 | 1.06-1.19 | 0.0002 |
| **Alc-23** |  | 1.15 | 1.09-1.22 | 1x10^-6^ |  | 1.15 | 1.08-1.22 | 4x10^-6^ |
| **Alc-5** |  | 1.14 | 1.08-1.21 | 4x10^-6^ |  | 1.14 | 1.07-1.21 | 1x10^-5^ |

^1^ RRs were calculated using conditional logistic regression models. Cases and controls were matched on age, sex, country of birth, sample type and smoking status (for the lung cancer study), Matched pairs were placed consecutively on a same chip of the assay, at random positions.

Model 1: adjusted for white blood cell composition

Model 2: additionally adjusted for smoking status (current/former/never), alcohol consumption (in g/day) and BMI (in kg/m2), smoking pack-years, age at starting (never smoked; age 16 or less; between age 17 and 21; after age 21 years); years since quitting (never smoked; more than 10 years without smoking, between 5 and 10 years without smoking; less than 5 years without smoking), a score for physical activity, the alternate healthy eating index to reflect overall diet quality, education (score ranging from 1 to 8), socioeconomic status (score ranging from 1 to 10) and height (continuous, in meters)
